# Appendix300: A multi-institutional laparoscopic appendectomy video dataset for computational modeling tasks

**DOI:** 10.1101/2025.09.05.25335174

**Authors:** Fiona R. Kolbinger, Max Kirchner, Kevin Pfeiffer, Sebastian Bodenstedt, Alexander C. Jenke, Julia Barthel, Matthias Carstens, Karolin Dehlke, Sophia Dietz, Sotirios Emmanouilidis, Guido Fitze, Martin Freitag, Fabian Holderried, Thorsten Jacobi, Weam Kanjo, Linda Leitermann, Sören Torge Mees, Steffen Pistorius, Conrad Prudlo, Astrid Seiberth, Jurek Schultz, Karolin Thiel, Daniel Ziehn, Stefanie Speidel, Jürgen Weitz, Jakob Nikolas Kather, Marius Distler, Oliver Lester Saldanha

## Abstract

**Background:** The limited availability of diverse and representative training data poses a critical barrier to the development of clinically relevant computational tools for intraoperative surgical decision support. Surgical procedures are not routinely recorded, and annotation requires domain expertise, resulting in a scarcity of open-access surgical video datasets with high-quality annotations. Existing datasets are typically limited to single institutions and specific procedures, such as cholecystectomy, and rarely comprise patient-level metadata like demographic characteristics, disease history, or laboratory parameters.

**Methods:** The Appendix300 dataset comprises 330 laparoscopic surgery recordings, including 325 full-length laparoscopic appendectomies and 5 control recordings from non-appendectomy procedures in pediatric and adult patients treated at five German centers. The dataset includes patient-level clinical metadata (demographics, medical history, clinical symptoms, laboratory parameters, and histopathological findings), as well as standardized expert annotations of the laparoscopic grade of appendicitis.

**Results:** Appendix300 currently represents the largest publicly available collection of surgical video data with patient metadata and the first curated dataset of laparoscopic appendectomies. It enables novel validation tasks for computer vision in surgery, including the classification of appendicitis severity and the detection of appendiceal perforation. Technical validation of the laparoscopic appendicitis grade annotations showed substantial interrater agreement (weighted Cohen’s κ = 0.615).

**Conclusion:** The Appendix300 dataset expands the scope of surgical data science by integrating video data with clinical and pathological metadata across institutions. It enables new and clinically relevant patient-level validation tasks for computer vision in laparoscopic surgery and facilitates decentralized learning approaches, overall enhancing the breadth and translational relevance of AI-based surgical video analysis.

**Dataset Description:** Appendix300 is a multi-institutional dataset comprising 330 laparoscopic surgery recordings, including 325 appendectomies and 5 control cases, detailed patient-level metadata (demographics, medical history, clinical symptoms, laboratory parameters, and histopathological findings), and expert annotations of appendicitis severity. It enables novel validation tasks for surgical AI, such as inflammation grading and perforation detection, and supports decentralized learning across diverse patient populations.

## Background & Summary

Computational analysis of medical imaging and clinical data has the potential to improve diagnostic accuracy and treatment stratification across medical fields. Increasing evidence from prospective clinical trials supports the clinical applicability and benefit of Artificial Intelligence (AI)-based tools.^1^ For example, computational tools have demonstrated expert-level performance for polyp detection during colonoscopy^2^, the detection of pulmonary nodules in chest X-rays^3^, and mammogram interpretation for breast cancer screening^4^.

Computational model performance and generalizability depend on the availability of large and diverse sets of clinical training and test data. Some clinical fields produce imaging as a means of documentation that becomes a component of patient records and is repetitively reviewed after acquisition for diagnostic follow-up or therapeutic decision-making (i.e., electrocardiograms in cardiology, X-rays in radiology, pictures of polyps in colonoscopy). In contrast, surgical video data is more complex to acquire and store in a standardized way due to its large file size. Reviewing and annotating surgical video data are time-consuming and require surgical domain knowledge.^5^ While surgical video recordings represent an objective documentation of a surgical procedure and capture information that could be used for educational and prognostic purposes^6–8^, surgeries are not routinely recorded, and retrospectively written operation notes remain the sole standard at most clinical institutions^9^. The largest available open-access datasets of intraoperative imaging comprise data from about 100 patients and cover a limited variety of procedures, including cholecystectomy (i.e., Cholec80^10^, CholecT50^11^) and cataract surgeries (i.e., Cataract-101^12^, CATARACTS^13^). Most datasets exclusively comprise spatial or categorical annotations of the surgical scene (i.e., presence of tools and anatomical structures in a video frame) rather than patient-level or clinical data, allowing for a limited number of surgical AI validation tasks that insufficiently represent the breadth of clinical reasoning tasks in surgery.^14^

To increase the diversity of annotated surgical video data and validation tasks for surgical data science^15^ approaches, we curated the Appendix300 dataset, comprising 330 video recordings of laparoscopic appendectomies and healthy control surgeries (i.e., appendix recordings from non-appendectomy laparoscopic surgeries) performed at five centers, as well as matched clinical metadata and annotations of the intraoperative grade of appendicitis (**Figure 1**). Appendix300 currently represents the largest publicly available collection of surgical video data with patient metadata and the first curated dataset of laparoscopic appendectomies. By combining patient-level clinical data and pathology-related annotations with laparoscopic video data, this dataset connects a range of structured data types along the diagnostic and therapeutic pathway, facilitating pathophysiology-related investigations. Furthermore, the multicentric nature of the dataset allows for the evaluation of decentralized learning approaches. In the context of acute appendicitis, potential future applications include the development of intraoperative assistance systems for quality control and a harmonization of the intraoperative inflammation grade and the histopathological phenotype, which are often discordant^16^. Relation with preoperative imaging^17^ and postoperative outcome data (i.e., surgical complications) are possible future extensions that are beyond the scope of this clinical use case but will need to be generated for other clinical use cases.

**Figure 1.**
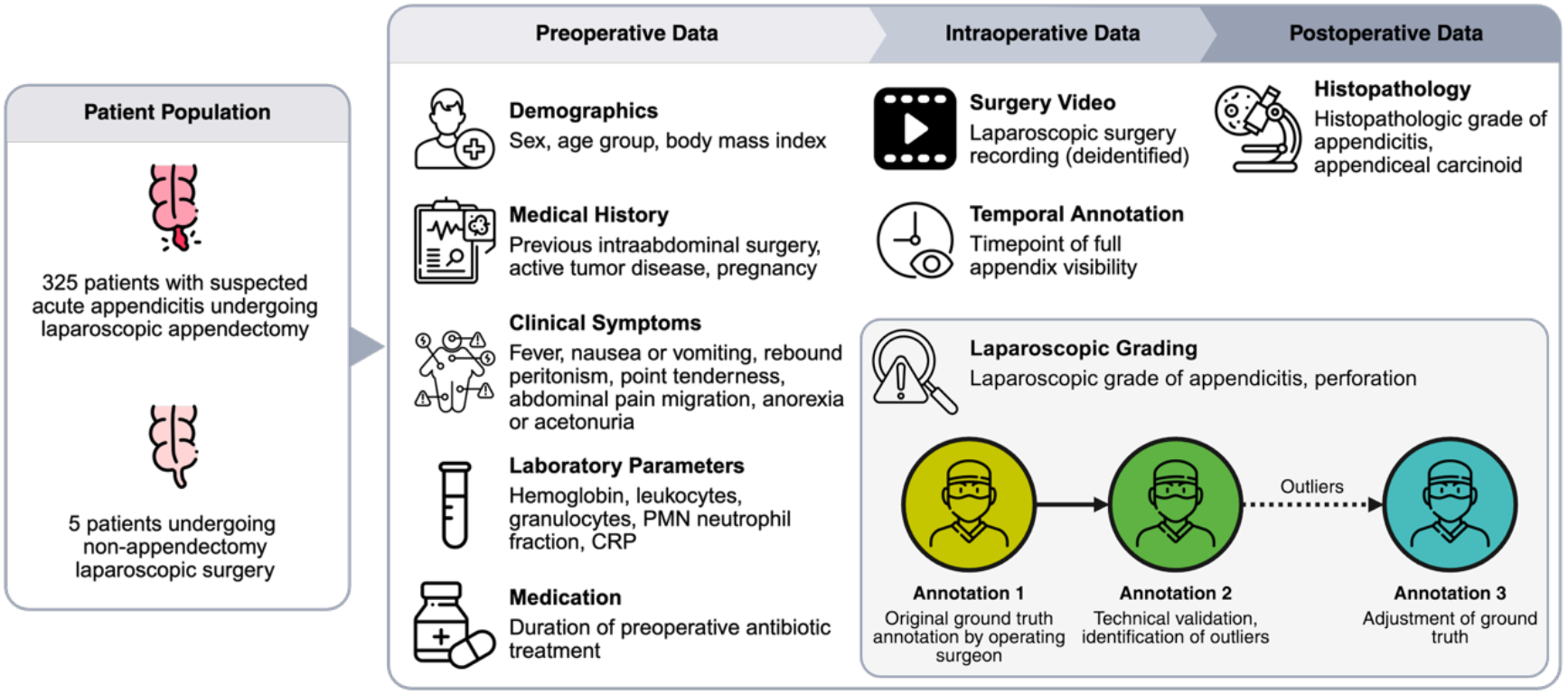
Dataset Overview. The Appendix300 dataset covers preoperative, intraoperative, and postoperative data of 325 patients with suspected appendicitis undergoing laparoscopic appendectomy and 5 patients undergoing non-appendectomy laparoscopic surgery at five surgical centers. Preoperative data comprise demographics, select medical history details, clinical symptoms of acute appendicitis, laboratory parameters, and recent antibiotic medication history. The surgery video recording was temporally annotated with regard to the timepoint of complete appendix visibility before appendix dissection, the laparoscopic grade of appendicitis, and perforation. Two surgeons independently annotated the laparoscopic grade and major disagreements were resolved through a third independent annotation. Postoperative metadata include the histopathological grade of appendicitis as well as the presence or absence of an appendiceal carcinoid. Abbreviations: CRP (C-reactive protein), PMN (polymorphonuclear).

## Methods

This dataset comprises laparoscopic video recordings of 325 laparoscopic appendectomies and 5 non-appendectomy laparoscopic surgeries, matched clinical metadata along the preoperative and postoperative treatment pathway, and annotations of the intraoperative grade of appendicitis (**Figure 1, Figure 2**).

**Figure 2.**
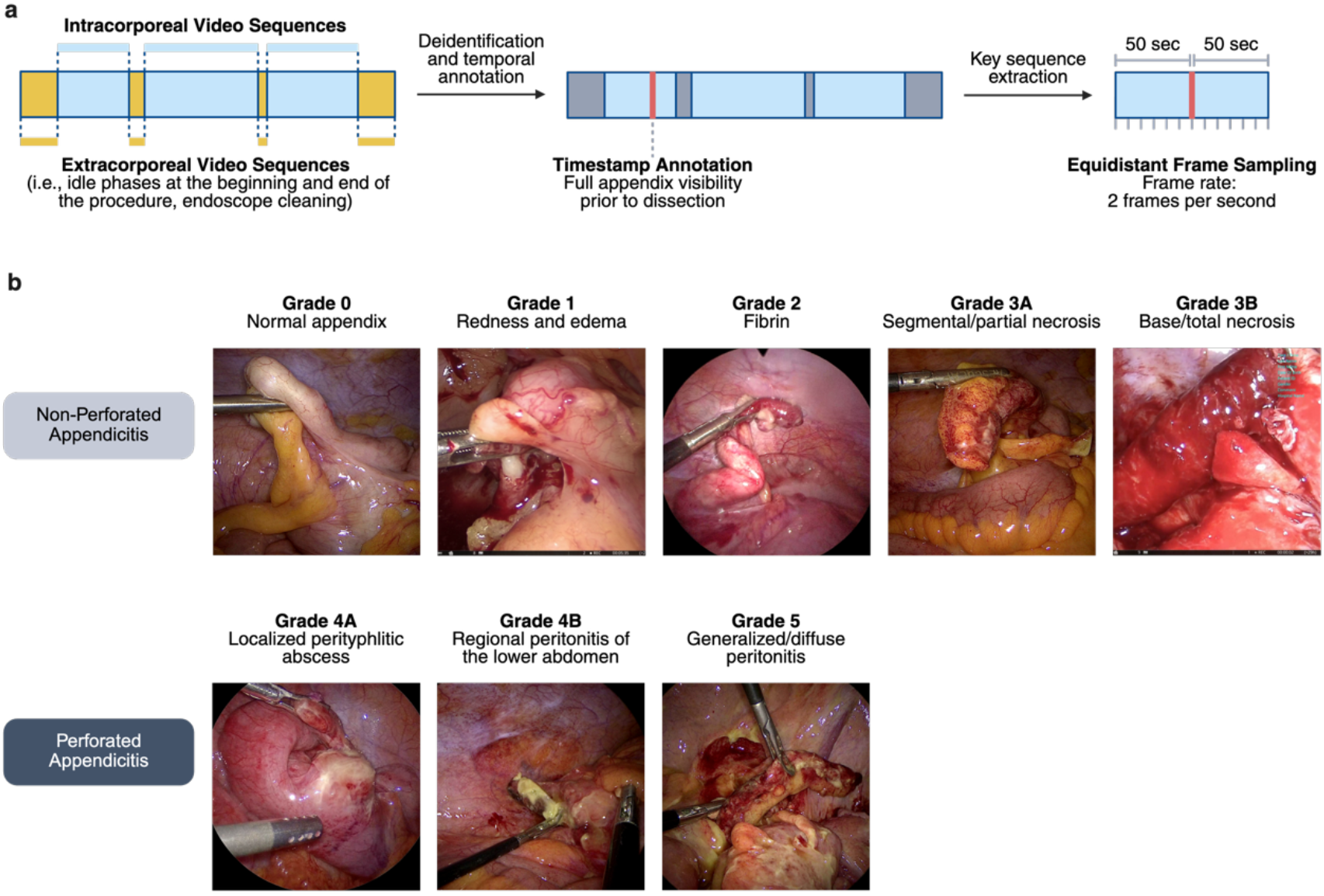
Data preprocessing and annotation of the intraoperative grade of appendicitis. **(a)** Schematic representation of the data processing steps. Following the deidentification of extracorporeal laparoscopic video sequences, each appendectomy recording was labeled with a temporal label (timestamp) marking the beginning of appendiceal dissection. Up to 200 equidistant frames were sampled from a 100-second key video sequence spanning 50 seconds before and after the timestamp. **(b)** Intraoperative grades of appendicitis were defined based on Gomes *et al*. (2012)^19^. Based on an annotation protocol (**Supplementary Material 3**), each video was classified by the surgical team. For technical validation, a second reviewer performed a second annotation of the intraoperative grade of appendicitis. In case of major disagreement, a third independent annotation was acquired to adjust the ground truth where necessary.

### Video Recording

Between June 2023 and September 2024, video data from 325 laparoscopic appendectomies and 5 non-appendectomy laparoscopic surgeries were gathered at five German surgical centers. The participating centers comprise four public hospitals (Asklepios-ASB Krankenhaus Radeberg, Diakonissenkrankenhaus Dresden, Krankenhaus St. Joseph Stift Dresden, St. Elisabethen-Krankenhaus Ravensburg) and one academic surgical department (Department of Pediatric Surgery, University Hospital Carl Gustav Carus Dresden). All included patients had a clinical indication for the surgical procedure.

Appendectomy recordings and clinical metadata were consecutively gathered in clinical routine. Patients with an indication of laparoscopic appendectomy for suspected acute appendicitis and documented histopathologic inflammation grade were included in the appendectomy dataset. Exclusion criteria comprised conversion to open surgery, incomplete surgery recordings, and cases with corrupted files. Surgeries were performed, recorded, and saved using locally available laparoscopic hardware (**Table 1**).

**Table 1:**
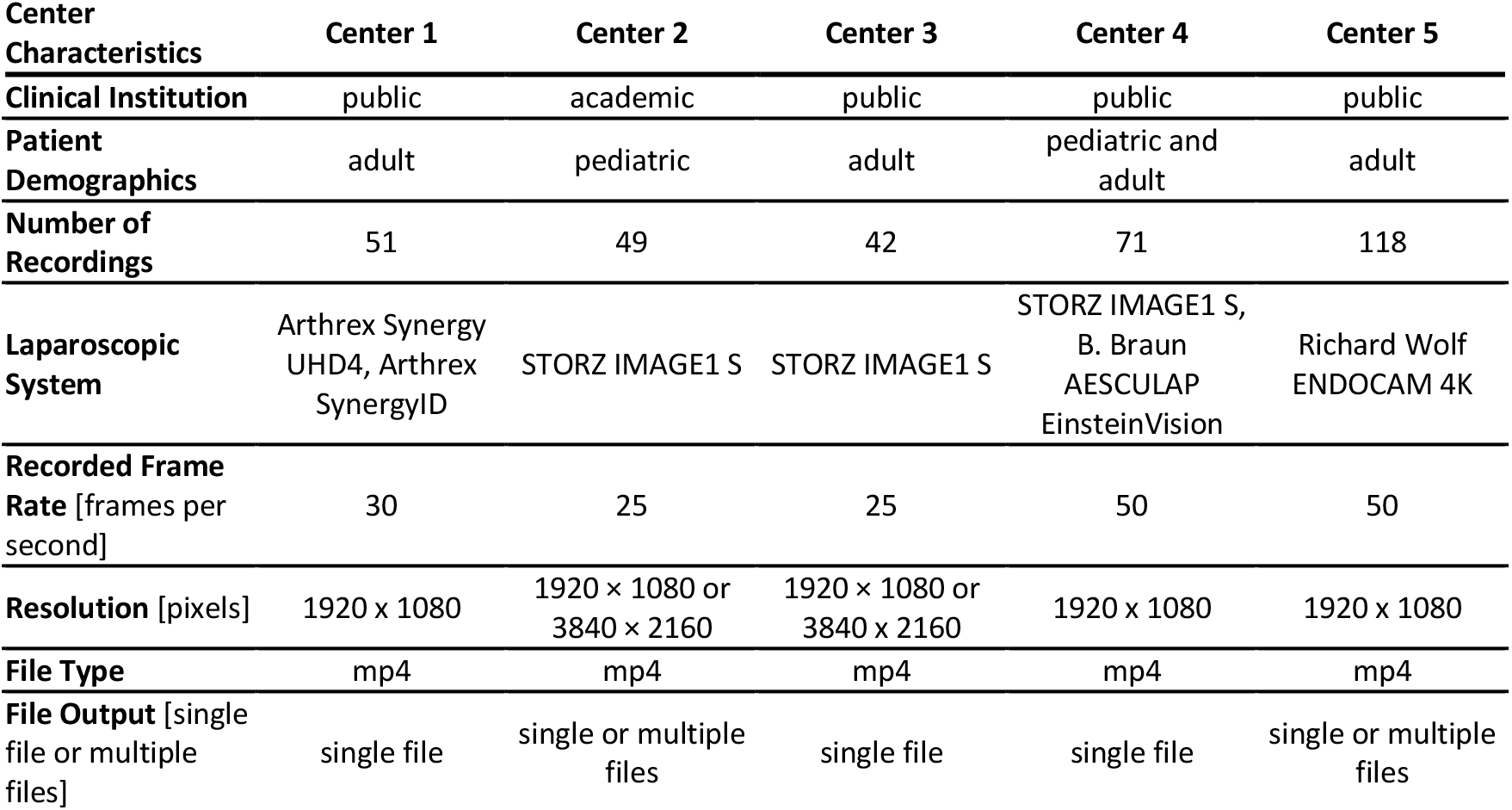
Recording infrastructure and technical details of the dataset for all contributing centers.

| Center Characteristics | Center 1 | Center 2 | Center 3 | Center 4 | Center 5 |
| --- | --- | --- | --- | --- | --- |
| **Clinical Institution** | public | academic | public | public | public |
| **Patient Demographics** | adult | pediatric | adult | pediatric and adult | adult |
| **Number of Recordings** | 51 | 49 | 42 | 71 | 118 |
| **Laparoscopic System** | Arthrex Synergy UHD4, Arthrex SynergyID | STORZ IMAGE1 S | STORZ IMAGE1 S | STORZ IMAGE1 S, B. Braun AESCULAP EinsteinVision | Richard Wolf ENDOCAM 4K |
| **Recorded Frame Rate** [frames per second] | 30 | 25 | 25 | 50 | 50 |
| **Resolution** [pixels] | 1920 x 1080 | 1920 x 1080 or 3840 x 2160 | 1920 x 1080 or 3840 x 2160 | 1920 x 1080 | 1920 x 1080 |
| **File Type** | mp4 | mp4 | mp4 | mp4 | mp4 |
| **File Output** [single file or multiple files] | single file | single or multiple files | single file | single file | single or multiple files |

### Acquisition of Clinical Labels and Annotations

A graphical user interface (**Supplementary Material 1**) was implemented to save raw surgery recordings, corresponding clinical labels, and original annotations of the laparoscopic grade of appendicitis on local hardware in anonymized form. This process was identical for laparoscopic appendectomy recordings and appendix recordings from non-appendectomy laparoscopic surgeries.

The graphical user interface was implemented in Python using tkinter and integrates a custom video playback component based on PyAV and Pillow for frame-accurate seeking, stereo video handling, and consistent frame rate rendering. It supports hospital-specific configurations (e.g., multi-video workflows, stereo cropping) via a centralized config structure and allows for timestamp annotation and structured metadata input through predefined form elements. All annotations and associated videos were saved to a defined output structure, with the metadata stored in a standardized CSV file, enabling reproducible and site-adaptive data extraction.

Clinical labels were derived from routine documentation based on a clinical labeling protocol (**Supplementary Material 2**). Following this protocol, the following parameters were preoperatively gathered: Sex, age, body-mass index (BMI), clinical symptoms based on the Alvarado score^18^ (migration of abdominal pain to the right lower quadrant, anorexia or acetone in the urine, nausea/vomiting, point tenderness in the right lower quadrant, rebound peritonism in the right iliac fossa, body temperature), preoperative laboratory parameters (hemoglobin, leukocytes, granulocytes, proportion of polymorphonuclear cells, C-reactive protein), select medical history details (history of intraabdominal surgery, active tumor disease, pregnancy), and the duration of preoperative antibiotic treatment (**Figure 1**).

Using the graphical user interface’s video review function (**Supplementary Material 1**), a timestamp, at which the appendix is fully visible prior to invasive preparation, was documented by the contributing surgery residents and attendings (FRK, JB, KD, SD, FH, WK, LL, JS, AS, DZ). Based on this timestamp, a 100-second video snippet was extracted from the full video recording, which covers 50 seconds before and after the timepoint of full appendix visibility (**Figure 2 a**).

The intraoperative grade of appendicitis and the presence of perforation were classified by the surgical teams carrying out the procedure based on an annotation protocol (**Supplementary Material 3, Figure 2 b**), which follows the intraoperative classification proposed by Gomes *et al*.^19^ (annotation 1). Annotation 1 represents the original ground truth annotation. For technical validation and to identify outliers and possible annotation errors, a second reviewer (general surgery resident with 4 years of experience in laparoscopic surgery) independently annotated the intraoperative grade of appendicitis for all recordings (annotation 2). Outliers were defined as a disagreement of at least three appendicitis grades between annotation 1 and annotation 2 (e.g., grade 2 and grade 5; grade 3A and grade 4B). We report the interrater agreement between annotation 1 and annotation 2 quantitatively using weighted Cohen’s kappa^20^ (**Table 2, Figure 3 a**), and qualitatively through a review of divergent ratings (**Figure 3 b**).

**Table 2:**
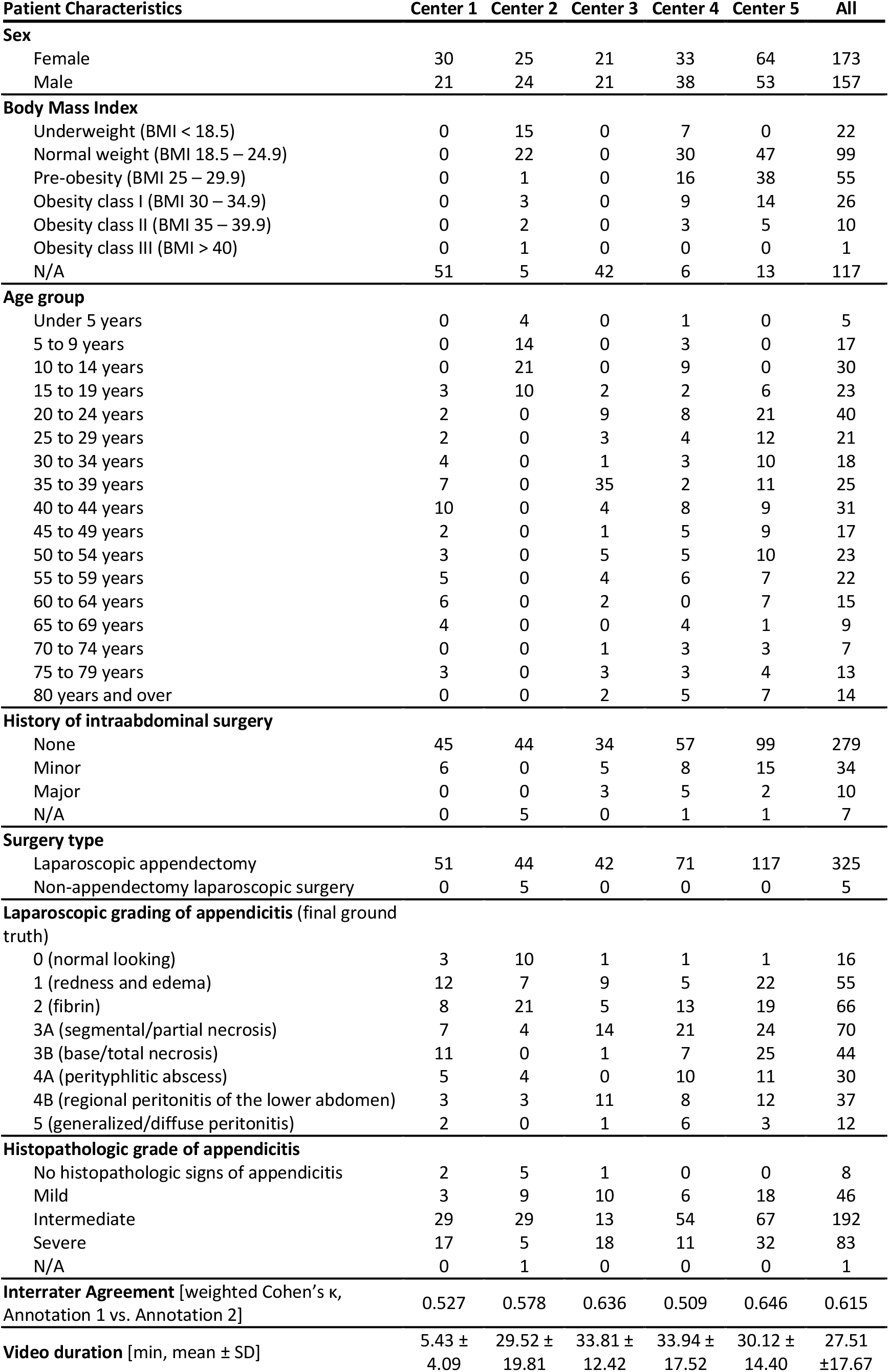
Summary of patient characteristics and surgery-related data.

**Figure 3.**
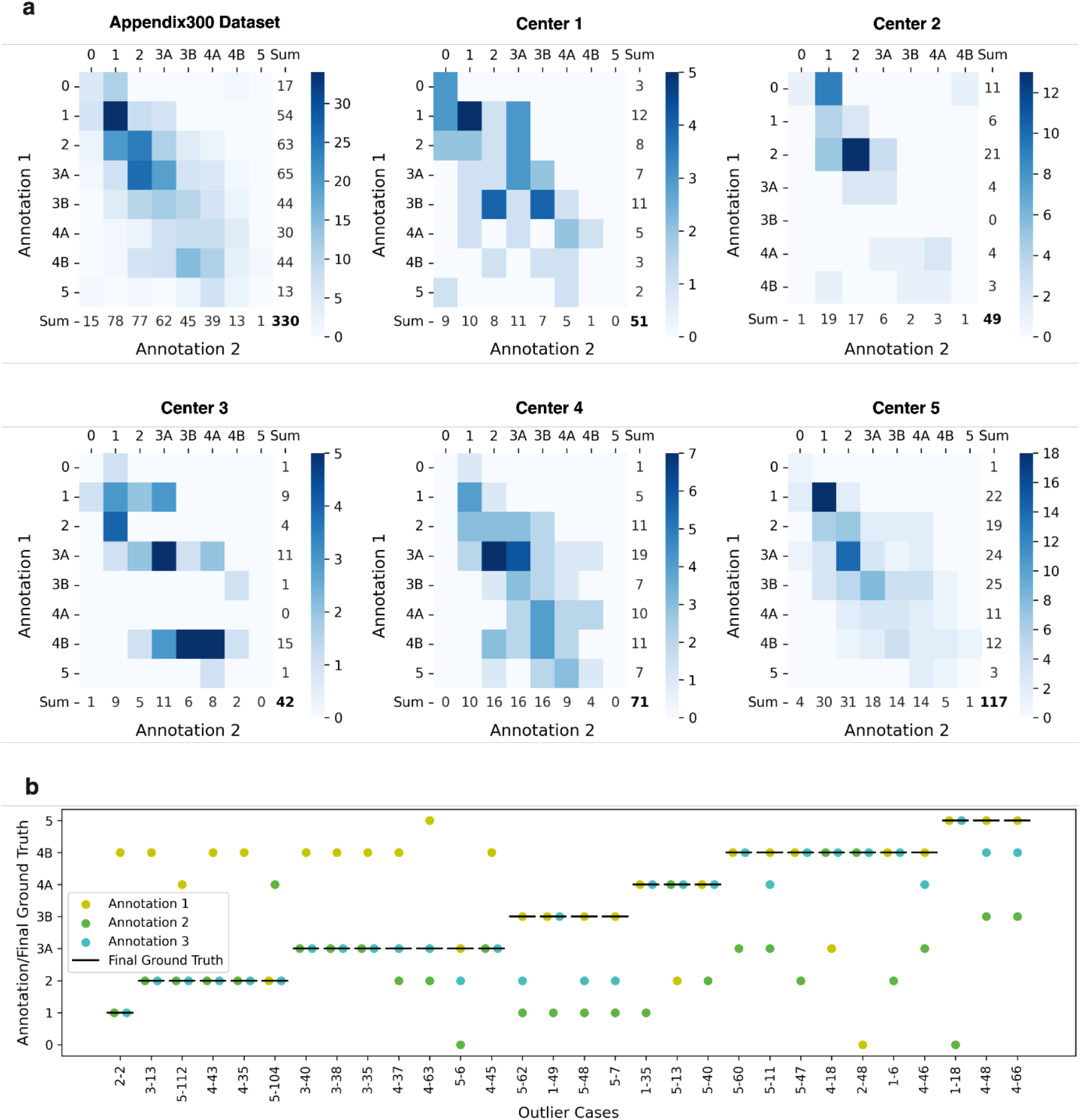
Technical validation of the laparoscopic grade of appendicitis annotations. Confusion matrices **(a)** indicate the agreement between the original annotation by the operating surgeon (annotation 1) and the independent second annotation (annotation 2) for all patients. **(b)** Of all 330 cases, 30 outlier cases with a disagreement of more than two appendicitis grades between annotation 1 and annotation 2 were subjected to a third independent annotation, and ground truth labels were adjusted as necessary. Case indices (x-axis ticks) comprise the center number and the case number, separated by a dash.

To adjust the ground truth where necessary, outliers were subjected to a third independent annotation by a general surgery resident with 7 years of experience in laparoscopic surgery (annotation 3). In case of a major disagreement between annotation 1 and annotation 3 (i.e., a disagreement of 3 or more appendicitis grades), the ground truth was adjusted to annotation 3. If annotation 3 was in agreement with annotation 1 (i.e., disagreement of up to 2 appendicitis grades), the ground truth was not adjusted (**Figure 3 b**).

**Table 2** summarizes patient characteristics and appendicitis grade annotations for all surgeries.

## Ethics Statement

This dataset was collected in accordance with the Declaration of Helsinki and its later amendments. All data were compiled in an anonymized fashion using the described user interface. The responsible Institutional Review Boards reviewed and approved this study on August 4th, 2022 (ethics committee at the Technical University Dresden, approval number BO-EK-332072022), September 13th, 2023 (ethics committee of the Sächsische Landesärztekammer, approval number EK-BR-75/23-1), and December 23rd, 2023 (ethics committee of the Landesärztekammer Baden-Württemberg, approval number B-F-2023-023). The trial, in the context of which this dataset was acquired, was prospectively registered at the German Clinical Trials Register (Deutsches Register Klinischer Studien, DRKS) on December 9th, 2022 (trial registration ID: DRKS00030874). Patients were informed about the anonymized acquisition, analysis, and publication of data from their inpatient treatment. Following local legislation, no written informed consent was required for anonymized acquisition, analysis, and publication of clinical data.

## Data Records

The Appendix300 dataset comprises 100-second video snippets from 330 surgery video recordings, along with patient metadata and annotations. These video snippets cover 50 seconds before and after the annotated timepoint of full appendix visibility. Due to the large file size of the deidentified full-length surgery recordings, they are made available to interested researchers upon reasonable request. To access the full-length surgery recordings, data requestors must sign a data use agreement.

During the peer review process, the data are accessible via the following link: https://nextcloud.tso.ukdd.de/s/mitAaKT8RiPqw3p. The dataset is publicly available for non-commercial use under the Creative Commons Attribution CC-BY. If readers wish to use or reference this dataset, they should cite this paper.

## Description of the data folder structure

The dataset is structured into two main directories: “Video_Snippets” and “Images”. Each of these directories is organized by contributing center, with data grouped into subfolders for each patient, such as “Center1_001”. The “Video_Snippets” directory contains patient subfolders with the respective 100-second video snippet covering 50 seconds before and after the annotated timepoint of full appendix visibility, following the same folder structure. In the “Images” directory, each patient subfolder includes a frames.zip archive with 200 frames extracted from the corresponding video snippet at 2 frames per second. At the top level of the dataset, an “overall_merged.csv” file is available to provide a consolidated view of all centers and patients, including metadata and annotations.

### Technical Validation

For technical validation of the annotations of the intraoperative grade of appendicitis, an independent secondary annotation was acquired. The overall interrater agreement (weighted Cohen’s kappa) was 0.614, with variations across the contributing centers (**Table 2, Figure 3 a**). Of 330 cases, 30 (9.1%) were classified as outliers or potential annotation errors based on a major disagreement between annotation 1 and annotation 2. Upon a third independent review of these 30 outlier cases (annotation 3), ground truths were adjusted to annotation 3 for 14 cases (4.2% of all cases) and original grade labels (annotation 1) were retained for 16 cases (**Figure 3 b**). These findings provide a baseline for interrater variability in laparoscopic grading of appendicitis.

### Usage Notes

The Appendix300 dataset can be used for various purposes in the field of machine learning, either on its own or in combination with other, already existing datasets. Used on its own, it facilitates the training of computational models identifying the intraoperative grade of appendicitis as a classification task and differentiation of perforated and non-perforated appendicitis (binary classification task), which are new clinical use cases for algorithm validation in surgical data science.

The multi-institutional nature of the Appendix300 dataset also facilitates the evaluation of decentralized and federated machine learning models for the above-mentioned validation tasks. We report the results of a benchmarking study evaluating Swarm Learning for decentralized, privacy-preserving collaboration in surgical video analysis in a separate publication.

The Appendix300 dataset has three key limitations: First, some clinical data are incompletely available due to the manual entry of anonymized metadata at contributing centers. For example, no information about the BMI could be obtained from center 1 and center 3 (**Table 2**). This limitation represents a common scenario in multi-institutional data collection efforts and could be overcome through direct access to electronic health records, which was not feasible in this work due to legal and administrative restrictions at the contributing centers. Second, the dataset does not include any preoperative imaging data or imaging-derived variables that have been identified as predictors of a complicated course, e.g., the presence of free intraabdominal air or an intra-abdominal abscess.^21^ Similarly, no data on postoperative complications are available in this dataset. Third, annotations of the laparoscopic grade of appendicitis followed the Gomes classification, which, in itself, has limitations.^19^ For example, this classification requires the presence of macroscopic necroses for the assignment of grade 3, and the presence of encapsulated abscesses or leakage of pus into the abdominal cavity for the assignment of grade 4 (**Figure 2 b, Supplementary Material 3**). While it is applicable to most laparoscopic presentations of appendicitis, some intermediate cases may be particularly ambiguous to classify. For example, an appendix presenting with fibrin coverage, mild regional peritonitis, and small amounts of opaque (i.e., likely purulent) ascites, yet without macroscopic necroses or encapsulated abscesses, may fall into either grade 2 (fibrin) or grade 4B (regional peritonitis). This ambiguity and potential non-linearity of the clinical progression of appendicitis and its laparoscopic presentation have been acknowledged by previous research.^22,23^

Despite these limitations, the Appendix300 dataset represents a considerable step towards clinically relevant applications for computational surgical video analysis, as it connects demographic, symptom-related, laparoscopic, and histopathological data for a large, multi-institutional patient cohort, and introduces a novel use case that will help diversify the clinical applications of research efforts in surgical data science.

## Data Availability

Data will be made publicly available following the peer review process for non-commercial use under the Creative Commons Attribution CC-BY. If readers wish to use or reference this dataset, they should cite this paper.

## Code Availability

No custom code was used in the generation or processing of this dataset.

## Acknowledgements

The authors gratefully acknowledge administrative support from Sandra Korn, Ulrike Neckmann, Christian Praetorius, and Anika Stützer (Clinical Trial Center, Department of Visceral, Thoracic and Vascular Surgery, University Hospital and Faculty of Medicine Carl Gustav Carus Dresden, Dresden, Germany) and technical support from all participating institutions in locally setting up data collection infrastructure.

This work was supported by the German Cancer Research Center (CoBot 2.0) and the German Research Foundation (Deutsche Forschungsgemeinschaft, DFG) as part of Germany’s Excellence Strategy (EXC 2050/1, Project ID 390696704) within the Cluster of Excellence “Centre for Tactile Internet with Human-in-the-Loop” (CeTI) of the Dresden University of Technology. FRK receives support from the Joachim Herz Foundation (Add-On Fellowship for Interdisciplinary Life Science), the Central Indiana Corporate Partnership AnalytiXIN Initiative, the Evan and Sue Ann Werling Pancreatic Cancer Research Fund, and the Indiana Clinical and Translational Sciences Institute (EPAR4157) funded, in part, by Grant Number UM1TR004402 from the National Institutes of Health, National Center for Advancing Translational Sciences, Clinical and Translational Sciences Award. MK and ACJ are supported by the European Union through NEARDATA under grant agreement ID 101092644. JNK is supported by the German Cancer Aid (DECADE, 70115166), the German Federal Ministry of Education and Research (PEARL, 01KD2104C; CAMINO, 01EO2101; SWAG, 01KD2215A; TRANSFORM LIVER, 031L0312A; TANGERINE, 01KT2302 through ERA-NET Transcan; Come2Data, 16DKZ2044A; DEEP-HCC, 031L0315A), the German Academic Exchange Service (SECAI, 57616814), the German Federal Joint Committee (TransplantKI, 01VSF21048) the European Union’s Horizon Europe and innovation programme (ODELIA, 101057091; GENIAL, 101096312), the European Research Council (ERC; NADIR, 101114631), the National Institutes of Health (EPICO, R01 CA263318) and the National Institute for Health and Care Research (NIHR, NIHR203331) Leeds Biomedical Research Centre. The views expressed are those of the author(s) and not necessarily those of the NHS, the National Institutes of Health, the NIHR, or the Department of Health and Social Care. This work was funded by the European Union. Views and opinions expressed are those of the authors only and do not necessarily reflect those of the European Union. Neither the European Union nor other granting authorities can be held responsible for them.

## Author Contributions

Conceptualization: FRK, JNK, MD, OLS; Methodology: FRK, SB, JS, SS, JNK, OLS; Software: MK, KP, SB, ACJ, OLS; Validation: FRK, MK, KP, SB, ACJ, MC, OLS; Formal analysis: FRK, MK, KP, OLS; Investigation: FRK, KP, SB, JB, MC, KD, SD, SE, FH, WK, LL, CP, AS, JS, DZ, MD, OLS; Resources: GF, MF, TJ, STM, SP, JS, KT, SS, JW, JNK, MD; Data Curation: FRK, MK, KP, SB, JB, MC, KD, SD, SE, FH, WK, LL, CP, AS, JS, DZ, MD, OLS; Writing - Original Draft: FRK; Writing - Review & Editing: MK, KP, SB, ACJ, JB, MC, KD, SD, SE, GF, MF, FH, TJ, WK, LL, STM, SP, CP, AS, JS, KT, DZ, SS, JW, JNK, MD; Visualization: FRK, MK; Supervision: FRK, SS, JW, JNK, MD, OLS; Project administration: FRK, OLS; Funding acquisition: FRK, SB, SS, JW, JNK, MD.

## Competing Interests

FRK declares advisory roles for Radical Healthcare, USA; and the Surgical Data Science Collective (SDSC), USA. JNK declares consulting services for Bioptimus, France; Owkin, France; DoMore Diagnostics, Norway; Panakeia, UK; AstraZeneca, UK; Scailyte, Switzerland; Mindpeak, Germany; and MultiplexDx, Slovakia. Furthermore, he holds shares in StratifAI, Synagen, and Ignition Labs, Germany, has received a research grant by GSK, and has received honoraria by AstraZeneca, Bayer, Eisai, Janssen, MSD, BMS, Roche, Pfizer and Fresenius. MD declares advisory roles for AESCULAP AG, Germany; and Intuitive Surgical, USA, and has received honoraria from Medtronic. All other authors declare no competing interests.

## Notes

### Clinical Protocols

https://drks.de/search/de/trial/DRKS00030874

### Author Declarations

This dataset was collected in accordance with the Declaration of Helsinki and its later amendments. All data were compiled in an anonymized fashion using the described user interface. The responsible Institutional Review Boards reviewed and approved this study on August 4th, 2022 (ethics committee at the Technical University Dresden, approval number BO-EK-332072022), September 13th, 2023 (ethics committee of the Saechsische Landesaerztekammer, approval number EK-BR-75/23-1), and December 23rd, 2023 (ethics committee of the Landesaerztekammer Baden-Wuerttemberg, approval number B-F-2023-023). The trial, in the context of which this dataset was acquired, was prospectively registered at the German Clinical Trials Register (Deutsches Register Klinischer Studien, DRKS) on December 9th, 2022 (trial registration ID: DRKS00030874). Patients were informed about the anonymized acquisition, analysis, and publication of data from their inpatient treatment. Following local legislation, no written informed consent was required for anonymized acquisition, analysis, and publication of clinical data.

### Summary of Updates

Correction of data distribution error

