## Supplementary Material for "Appendix300: A multi-institutional laparoscopic appendectomy video dataset for computational modeling tasks"

### Supplementary Material 1: Graphical User Interface

**a**

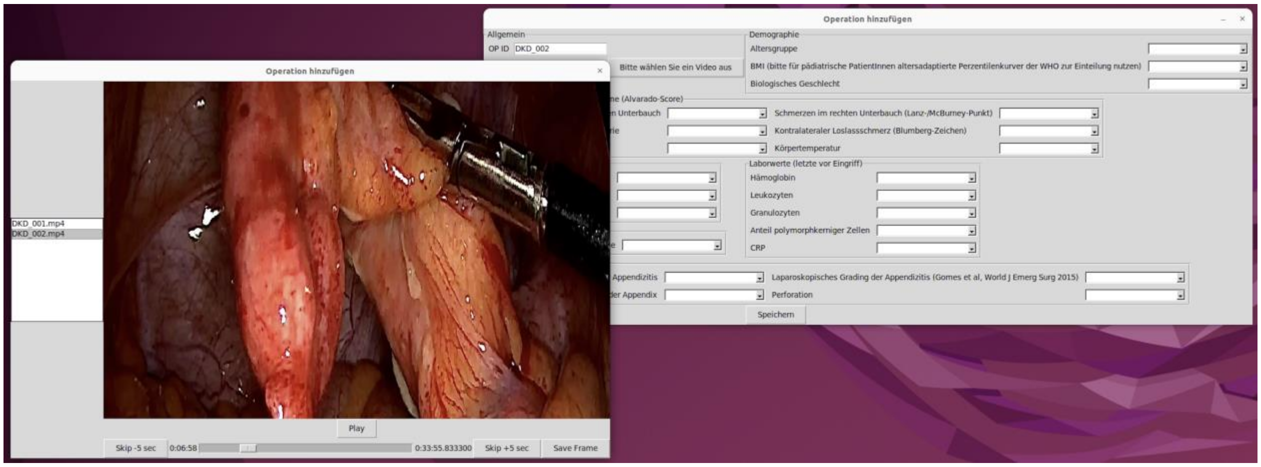

**b**

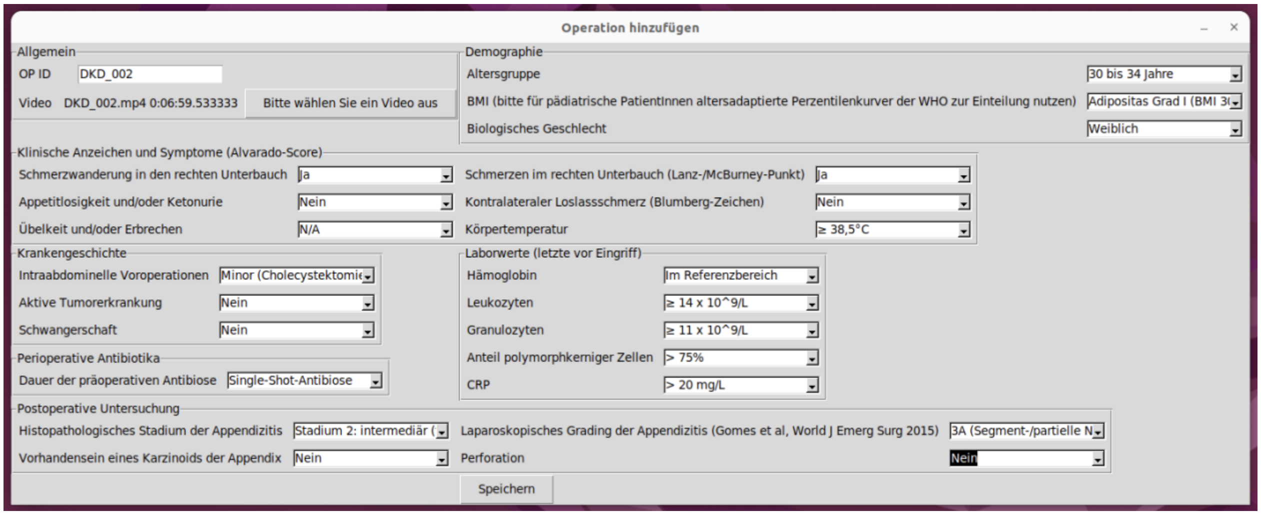

**Supplementary Material 1: Graphical user interface for video and clinical data collection and annotation.** The graphical user interface facilitates the upload and review of raw surgery recordings to annotate the timepoint of full appendix visibility before appendiceal dissection **(a)**. Subsequently, corresponding clinical labels and original annotations of the laparoscopic grade of appendicitis were entered at contributing centers **(b)**. To guarantee data privacy and minimize annotation errors, all clinical data were entered in categorical format using a dropdown menu. The graphical user interface was provided to contributing centers in English and German.

### Supplementary Material 2: Clinical Labeling Protocol

**Demographic parameters:**

- **Age group**: Age as a categorical variable, in 5-year intervals:
  - < 5 years
  - 5-9 years
  - 10-14 years
  - […]
  - 75-79 years
  - > 80 years
- **BMI**: Body Mass Index as a categorical variable according to the World Health Organization (WHO) definition. For pediatric patients and adolescents up to 20 years of age, BMI was categorized using age-adapted percentile curves, according to the WHO definition[1]:
  - Underweight (BMI < 18,5)
  - Normal weight (BMI 18,5 – 24,9)
  - Pre-obesity (BMI 25 – 29,9)
  - Obesity class I (BMI 30 – 34,9)
  - Obesity class II (BMI 35 – 39,9)
  - Obesity class III (BMI > 40)
- **Sex**: Biological sex
  - Male
  - Female
  - N/A

**Clinical symptoms**: Binary documentation based on the Alvarado score[2]:

- **Migration of abdominal pain to the right lower quadrant** – *German: Schmerzwanderung in den rechten Unterbauch*
  - Yes
  - No
  - N/A
- **Anorexia (or acetone in the urine)** – *German: Appetitlosigkeit oder Ketonurie*
  - Yes
  - No
  - N/A
- **Nausea/vomiting** – *German: Übelkeit/Erbrechen*
  - Yes
  - No
  - N/A
- **Point Tenderness in the right lower quadrant** – *German: Schmerzen im rechten Unterbauch (Lanz-/McBurney-Punkt)*
  - Yes
  - No
  - N/A

- **Rebound peritonism in the right iliac fossa** – *German: Kontralateraler Loslassschmerz (Blumberg-Zeichen)*
  - Yes
  - No
  - N/A
- **Body temperature** – *German: Körpertemperatur*
  - < 37.7°C
  - ≥ 37.7°C
  - ≥ 38.5°C
  - N/A

**Clinical history:**

- **History of intraabdominal surgery**: Prior intraabdominal surgeries, documented as a categorical variable:
  - None
  - Minor (cholecystectomy, cesarean section)
  - Major
- **Active tumor disease**: Binary documentation of any active tumor disease at the time of surgery
  - Yes
  - No
  - N/A
- **Pregnancy**: Binary documentation of pregnancy
  - Yes
  - No
  - N/A (male patient)
- **Duration of preoperative antibiotic treatment**, documented as a categorical variable:
  - no preoperative antibiosis
  - single-shot antibiosis
  - 1 day
  - 2 days
  - 3 days
  - 4 days
  - 5 days
  - 6 days
  - ≥ 7 days
  - N/A

**Laboratory parameters**: Based on the last set of laboratory parameters before surgery, documented as a categorical variable[3]:

- **Hemoglobin**
  - In reference range

- Below reference range
- Above reference range
- N/A

- **Leukocytes**
  - $< 10 \times 10^9/L$
  - $\geq 10 \times 10^9/L$
  - $\geq 12 \times 10^9/L$
  - $\geq 14 \times 10^9/L$
  - $\geq 15 \times 10^9/L$
  - N/A
- **Granulocytes**
  - $< 7 \times 10^9/L$
  - $\geq 7 \times 10^9/L$
  - $\geq 9 \times 10^9/L$
  - $\geq 11 \times 10^9/L$
  - $\geq 13 \times 10^9/L$
  - N/A
- **Proportion of polymorphonuclear cells**
  - ≤ 75%
  - > 75%
  - > 85%
  - N/A
- **C-reactive protein**
  - ≤ 10 mg/L
  - > 10 mg/L
  - > 20 mg/L
  - N/A

**Histopathology:**

- **Histopathologic grade of appendicitis**: Several different classifications exist for histopathological assessment of acute appendicitis. In clinical practice, appendectomy specimens are often described inconsistently and not necessarily following any published classification. Therefore, the following classification[4] was used in this study:
  - Grade 1: mild, i.e., mild inflammation restricted to mucosa
    *Common German terms: "auf die Mukosa begrenzt", "erosiv", "katarrhalisch"*
  - Grade 2: intermediate, i.e., transmural inflammation
    *Common German terms: "transmural", "phlegmonös", "ulcerophlegmonös"*
  - Grade 3: severe
    *Common German terms: “perforiert", "gangränös"*
- **Appendiceal carcinoid**: Presence of an appendiceal carcinoid based on the final pathology report:
  - Yes
  - No
  - N/A

#### Supplementary Material 3: Video Acquisition and Annotation Protocol

For maximum clarity, the video acquisition and annotation protocol was provided in English and German.

##### Video Acquisition

**Laparoscopic appendectomy**

- Start recording at insertion of the camera into the abdomen
- End recording at the end of the surgery, when the laparoscope is removed from the abdomen
- Do not interrupt the recording when camera is removed from the abdomen (e.g., to clean the lens)

**Laparoskopische Appendektomie**

- Beginn der Aufzeichnung bei Einführung der Kamera in das Pneumoperitoneum
- Ende der Aufzeichnung bei Entfernung der Kamera aus dem Pneumoperitoneum nach Operationsende
- Keine Unterbrechung der Aufnahme bei Entfernung der Kamera aus dem Pneumoperitoneum (z.B. zur Reinigung der Kamera)

**Laparoscopic right hemicolectomy**

- Record a part of the surgery, in which you expose the appendix
- Duration of the video: at least 1 minute
- Start recording before starting to expose the appendix
- End recording after exposure of the appendix is complete
- Include a view with the appendix fully visible until its base, similar to the process during an appendectomy

- If possible, please order a specific histopathological examination of the appendix when sending the specimen for histopathological review

**Laparoskopische Hemikolektomie rechts**

- Aufzeichnung eines Abschnitts der Operation, in dem die Appendix freipräpariert und exponiert wird
- Dauer des Videoausschnitts: mindestens 1 Minute
- Beginn der Aufzeichnung vor Beginn der Appendix-Präparation
- Ende der Aufzeichnung nach Ende der Appendix-Exposition
- Aufnahme einer Ansicht, in der die Appendix vollständig freipräpariert und exponiert ist (ähnlich dem Vorgehen bei einer Appendektomie)

- Wenn möglich, bitte spezifische histopathologische Begutachtung der Appendix bei Einsendung des Hemikolektomie-Präparats

#### Intraoperative Assessment of Appendicitis (based on Gomes *et al.*, 2012[5])

##### Grade 0: Normal looking appendix

— Normal appendix
— No relevant signs of inflammation
— No significant hyperemia

##### Grad 0: Normale Appendix

— Normale Appendix
— Keine relevanten Entzündungszeichen
— Keine relevante Gefäßinjektion

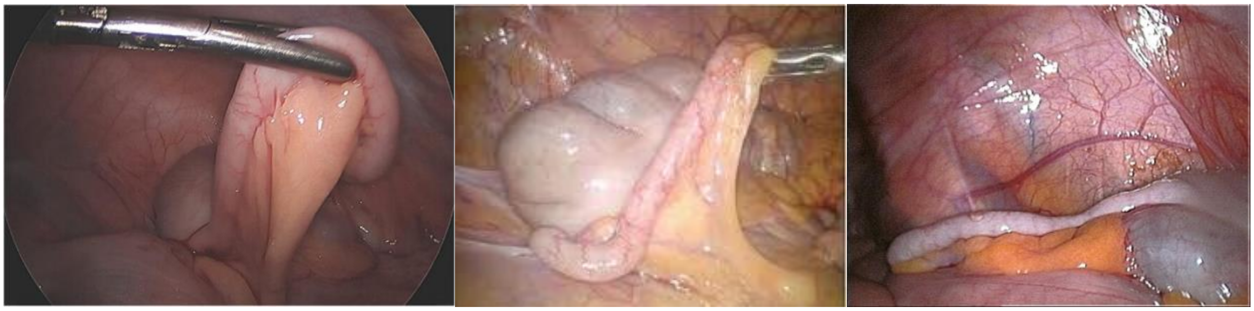

##### Grade 1: Redness and edema

— Redness and subtle edematous swelling of the appendix
— Hyperemia
— Likely corresponds to reversible phase of appendicitis

##### Grad 1: Rötung und Ödem

*Synonym: Katarrhalische Appendizitis*

— Rötung und leichte Ödembildung der Appendix
— Relevante Gefäßinjektion
— Entspricht a.e. reversibler Phase der Appendizitis

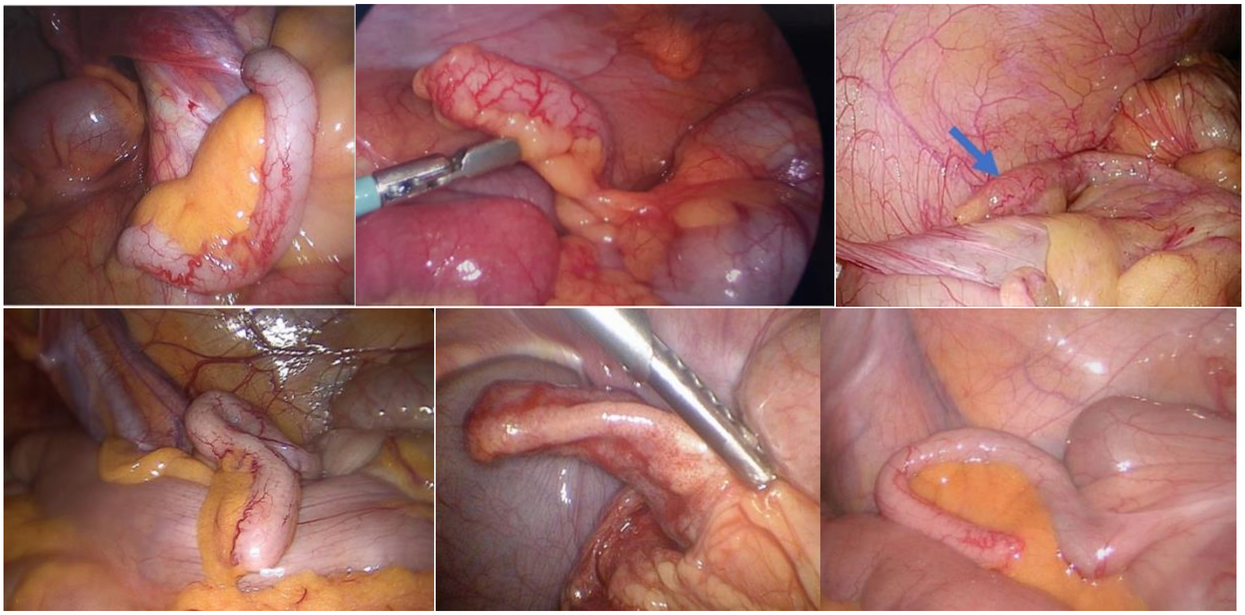

**Grade 2: Fibrin**

— Significantly increased size/diameter of the appendix
— Fibrinous exsudation
— No necrosis, no perforation
— Likely corresponds to transmural infection

**Grad 2: Fibrinexsudation**
*Synonym: (Sero-)Purulente Appendizitis, phlegmonöse Appendizitis*

— Deutliche Vergrößerung der Appendix in Länge und Durchmesser
— Fibrinexsudation
— Keine Nekrosen, keine Perforation
— Entspricht a.e. transmuraler Entzündung

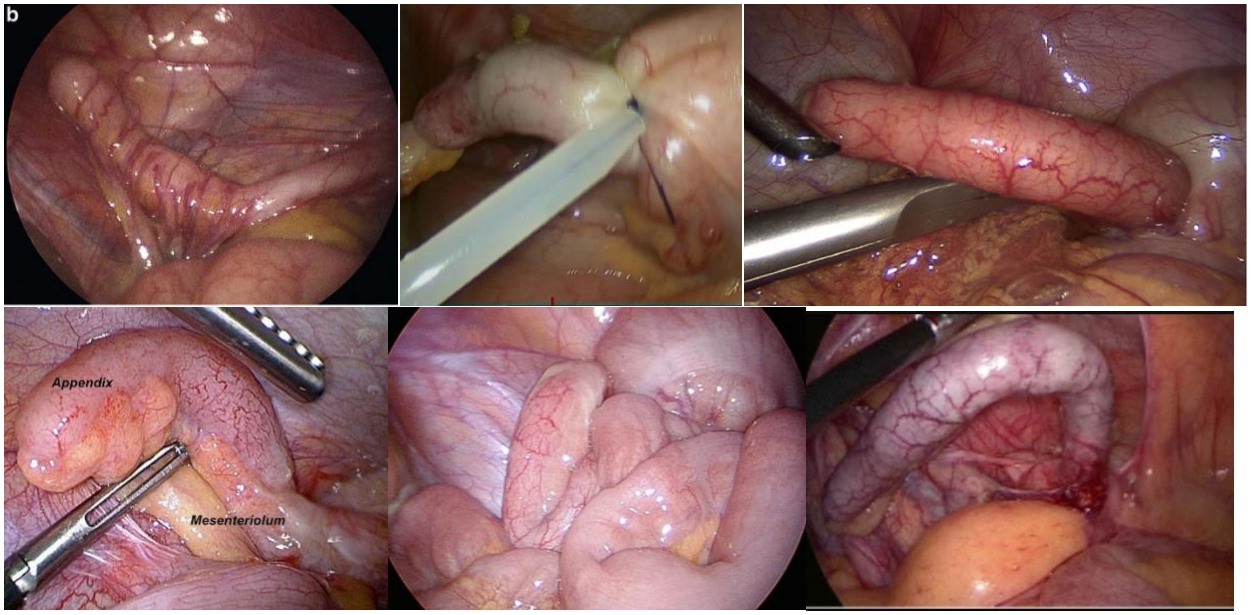

**Grade 3A: Segmental/partial necrosis**

— Localized green, brown or black discoloration
— Fibrinous exsudation
— No necroses at the appendiceal base, no perforation
— Likely corresponds to transmural infection, risk of perforation when manipulating the appendix

**Grad 3A: Segmentelle/partielle Nekrosen**
*Synonym: Segmentelle gangränöse Appendizitis*

— Stellenweise livide, grünliche, bräunliche oder schwarze Verfärbung der Appendix
— Fibrinexsudation
— Keine Nekrosen an der Basis, keine Perforation
— Entspricht a.e. transmuraler Entzündung, Perforationsgefahr bei Manipulation

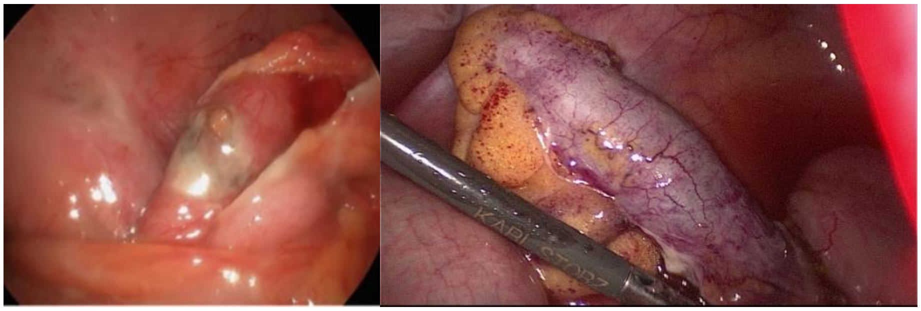

**Grade 3B: Base/total necrosis**

- Green, brown or black discoloration at the appendiceal base or of the entire appendix
- Fibrinous exsudation
- No perforation
- Likely corresponds to transmural infection, risk of perforation when manipulating the appendix

**Grad 3B: Basisnekrose/vollständige Nekrose**
*Synonym: Vollständige gangränöse Appendizitis*

- Livide, grünliche, bräunliche oder schwarze Verfärbung der Appendixbasis oder der kompletten Appendix
- Fibrinexsudation
- Keine Perforation
- Entspricht a.e. transmuraler Entzündung, Perforationsgefahr bei Manipulation

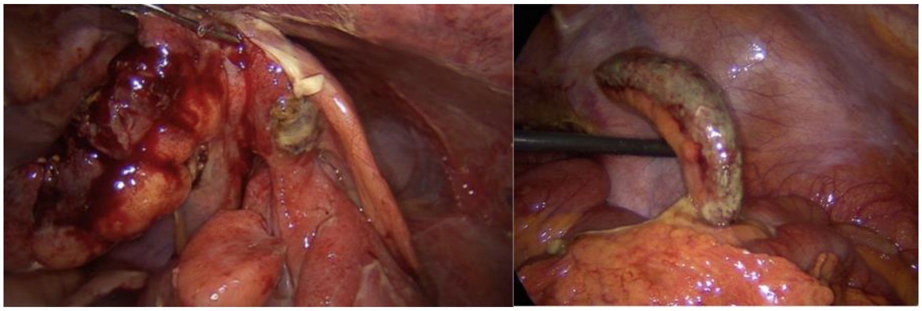

**Grade 4A: Localized perityphlitic abscess**

- Perforated appendix with localized (i.e. retroperitoneal) leakage, abscess covered by other tissue
- No "free" leakage of pus/intestinal content into the abdominal cavity
- Likely corresponds to transmural infection with perforation

**Grad 4A: Lokalisierter perityphlitischer Abszess**
*Synonym: Gedeckte Perforation mit perityphlitischem Abszess/Periappendizitis*

- Perforation der Appendix mit lokalisiertem Flüssigkeitsaustritt (z.B. retroperitoneal), Abszess gedeckt
- Keine „freie" Leckage von Eiter/Darminhalt in die Bauchhöhle
- Entspricht a.e. transmuraler Entzündung mit Perforation

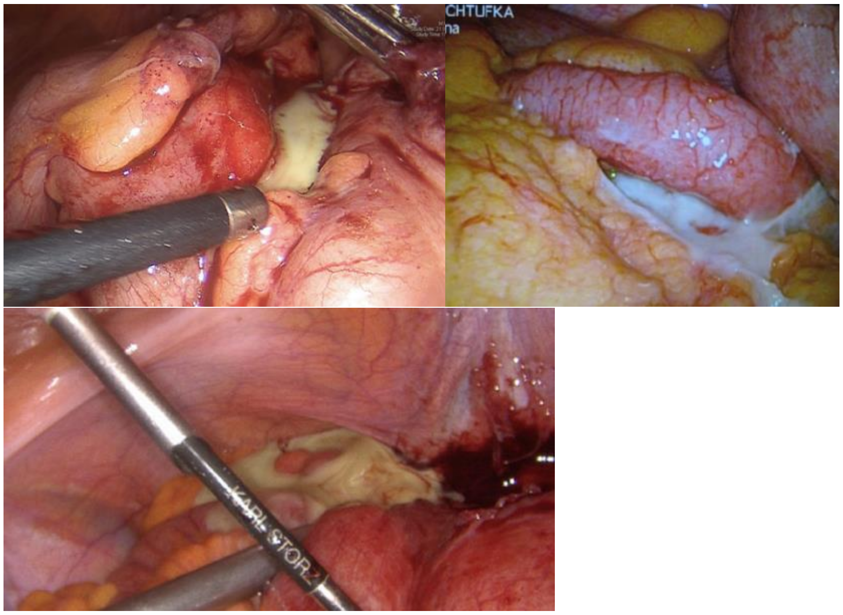

**Grade 4B: Regional peritonitis of the lower abdomen**

- Perforated appendix with leakage of pus/intestinal content into the abdominal cavity
- No encapsulated abscess (abscess not covered by other tissue)
- Regional peritonitis of the lower (not the upper) abdominal quadrants
- Likely corresponds to transmural infection with perforation

**Grad 4B: Regionäre Peritonitis des Unterbauchs**
*Synonym: Freie Perforation mit lokaler Unterbauch-Peritonitis*

- Perforation der Appendix mit freiem Austritt von Eiter/Darminhalt in die Bauchhöhle
- Kein abgekapselter/gedeckter Abszess
- Regionäre Peritonitis (nur im Unter-, nicht im Oberbauch)
- Entspricht a.e. transmuraler Entzündung mit Perforation

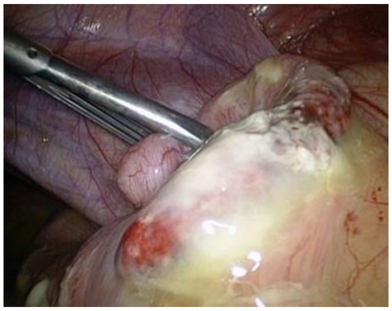

**Grade 5: Generalized/diffuse peritonitis**

- Perforated appendix with leakage of pus/intestinal content into the abdominal cavity
- No encapsulated abscess (abscess not covered by other tissue)
- Peritonitis of all abdominal quadrants
- Likely corresponds to transmural infection with perforation

**Grad 5: Generalisierte/diffuse Peritonitis**
*Synonym: Freie Perforation mit diffuser Peritonitis*

- Perforation der Appendix mit freiem Austritt von Eiter/Darminhalt in die Bauchhöhle
- Kein abgekapselter/gedeckter Abszess
- Vier-Quadranten-Peritonitis
- Entspricht a.e. transmuraler Entzündung mit Perforation

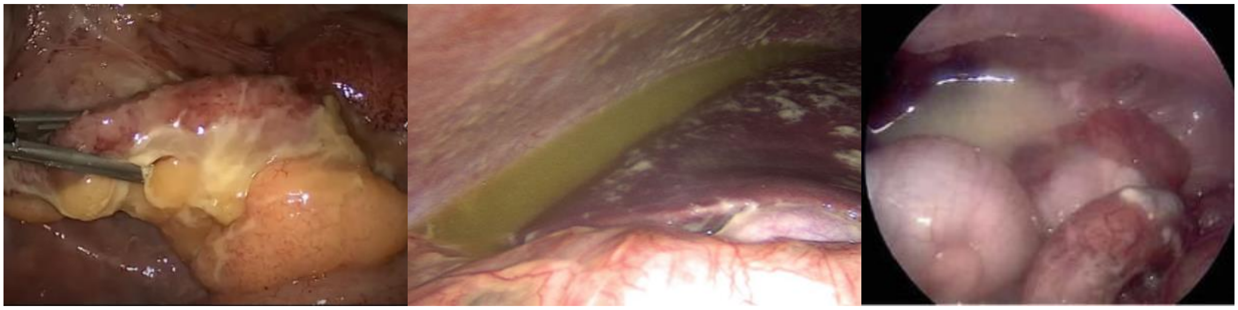
